# Evaluating the Performance of Artificial Intelligence in Generating Differential Diagnoses for Infectious Diseases Cases: A Comparative Study of Large Language Models

**DOI:** 10.1101/2024.06.28.24309694

**Authors:** Agnibho Mondal, Rucha Karad, Boudhayan Bhattacharjee, Bibhuti Saha

**Affiliations:** Department of Infectious Diseases and Advanced Microbiology, School of Tropical Medicine, Kolkata

## Abstract

**Background:** Artificial Intelligence (AI) has potential to transform healthcare including the field of infectious diseases diagnostics. This study assesses the capability of three large language models (LLMs), GPT 4, Llama 3, and Gemini 1.5 to generate differential diagnoses, comparing their outputs against those of medical experts to evaluate AI’s potential in augmenting clinical decision-making.

**Methods:** This study evaluates the differential diagnosis capabilities of three LLMs, GPT 4, Llama 3, and Gemini 1.5, using 50 simulated infectious disease cases. The cases were diverse, complex, and reflective of common clinical scenarios, including detailed histories, symptoms, lab results, and imaging findings. Each model received standardized case information and produced differential diagnoses, which were then compared to reference differential diagnosis lists created by medical experts. The analysis utilized the Jaccard index and Kendall’s Tau to assess similarity and order accuracy, summarizing findings with mean, standard deviation, and combined p-values.

**Results:** The mean numbers of differential diagnoses generated by GPT 4, Llama 3, and Gemini 1.5 were 6.22, 5.06, and 10.02 respectively which was significantly different (p*<*0.001) from the medical experts. The mean Jaccard index of GPT 4, Llama 3, and Gemini 1.5 were 0.3, 0.21, and 0.24 while the mean Kendall’s Tau were 0.4, 0.7, and 0.33 respectively. The combined p-value of GPT 4, Llama 3, and Gemini 1.5 were 1, 1, 0.979 respectively indicating no significant association between the differential diagnosis generated by the LLMs and the medical experts.

**Conclusion:** Although LLMs like GPT 4, Llama 3, and Gemini 1.5 exhibit varying effectiveness, none align significantly with expert-level diagnostic accuracy, emphasizing the need for further development and refinement. The findings highlight the importance of rigorous validation, ethical considerations, and seamless integration into clinical workflows to ensure AI tools enhance healthcare delivery and patient outcomes effectively.

## Introduction

Artificial Intelligence (AI) has rapidly transformed numerous sectors, and healthcare is no exception. Over recent years, AI’s integration into medical diagnostics has shown promising advancements, particularly in the field of infectious diseases. The ability of AI to analyze vast amounts of data and generate accurate differential diagnoses holds great potential to augment clinical decision-making and improve patient outcomes.

AI applications in healthcare encompass various domains, including predictive analytics, image analysis, and natural language processing (NLP). These technologies have been used to predict sepsis in high-risk patients, analyze host-response proteomic data to forecast respiratory infections, and identify pathogens in clinical samples.[1] Furthermore, AI models have been instrumental in enhancing the diagnostic processes during the COVID-19 pandemic by accurately identifying and predicting disease severity.[2, 3]

The use of large language models (LLMs) such as GPT 4, Llama 3, and Gemini 1.5 in generating differential diagnoses represents a significant leap in AI capabilities. While GPT 4 and Gemini 1.5 are proprietary LLMs developed by OpenAI and Google respectively, Llama 3 is an open-source model developed by Meta. These models leverage vast training datasets and advanced algorithms to understand and process medical information, thereby aiding in the diagnostic process. Prior studies have highlighted the efficacy of machine learning (ML) and AI in infectious disease diagnostics, especially in immunocompromised patients who are at a higher risk for severe infections.[4]

AI and ML are transforming various fields, including medicine and infectious diseases, with a particular focus on explainable AI/ML models for better understanding and managing these diseases.[5] AI advancements could automate image analysis for pathogen identification and classification of colony growth to enhance diagnostic accuracy and efficiency.[6] ML is also being increasingly used to predict antibiotic resistance based on pathogen genomes which could help in controlling antimicrobial resistance.[7]

This study aims to compare the differential diagnosis capabilities of three prominent LLMs including GPT 4, Llama 3, and Gemini 1.5. By examining the performance of these LLMs, we seek to understand their strengths and limitations in clinical settings and explore their potential integration into healthcare practices. This comparative analysis not only contributes to the body of knowledge on AI in medical diagnostics but also highlights the evolving role of AI in enhancing the accuracy and efficiency of healthcare delivery.

## Methodology

This study compares the differential diagnosis capabilities of three LLMs including GPT 4, Llama 3, and Gemini 1.5. For this comparison, simulated cases, rather than real world patient data, were used due to ethical implications.

To evaluate these models, we curated 50 simulated infectious disease cases. These cases were chosen to encompass a wide range of infectious diseases. The selection criteria included diversity, ensuring a broad spectrum of diseases; complexity, including cases with varying levels of diagnostic difficulty; and relevance, selecting cases commonly encountered in clinical practice. The cases were designed to include detailed patient histories, symptoms, laboratory results, and imaging findings, structured to mimic real-world clinical scenarios.

Each LLM was provided with the 50 simulated cases in a standardized input format, which included all relevant clinical details necessary for generating a list of differential diagnoses. The process involved input standardization, ensuring consistent case presentation to the models; prompting the models with a structured clinical query; and collecting the models’ outputs in a formatted manner for analysis.

The AI-generated differential diagnoses were benchmarked against reference lists formulated by three medical experts. The experts were provided with the same 50 simulated cases and asked to generate reference lists of differential diagnoses for each case by consensus. Their methodology involved a thorough review of each case, generating a list of potential diagnoses in the order of likelihood based on clinical judgment and expertise.

The lists generated by the LLMs were then compared against the reference list. The similarity was calculated using Jaccard index. Additionally, Kendall’s Tau was calculated to take the ordering of the differential diagnosis into account. The Jaccard index, Kendall’s Tau and p-value were calculated for each case. Overall summarization was done by calculating mean and standard deviation (SD) of Jaccard index and Kendall’s Tau while the combined p-value was calculated using Fisher’s combined probability method.

Statistical analysis was done using Python 3.12.3 and R 4.4.0 by R Foundation for Statistical Computing. A p-value less than 0.05 was considered significant.

## Results

A total number of 50 simulated infectious diseases cases were included in the study. The mean number of differential diagnoses generated by the medical experts was 8.48 while GPT 4, Llama 3, and Gemini 1.5 generated a mean number of 6.22, 5.06, and 10.02 differential diagnoses respectively as shown in figure 1. The count of differential diagnosis was significantly different (p<0.001) between the expert and the LLMs as well as between each LLM themselves.

**Figure 1:**
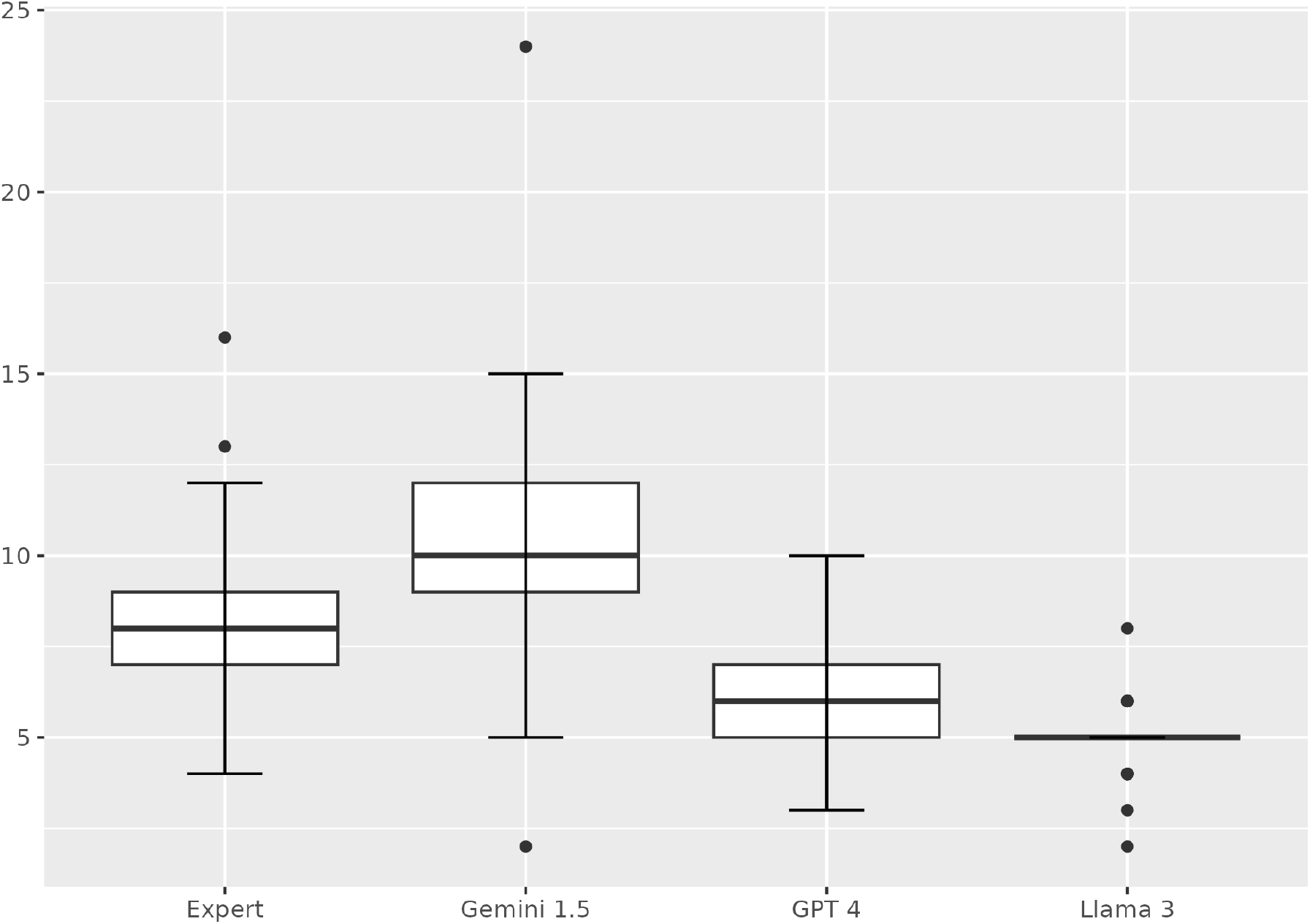
Comparison of the number of differential diagnoses generated by the medical experts and the large language models

The mean Jaccard index of GPT 4 was 0.3 (SD 0.17). The mean Jaccard index for Llama 3 was 0.21 (SD 0.13) while Gemini 1.5 had a mean Jaccard index of 0.24 (SD 0.11).

The mean Kendall’s Tau for GPT 4, Llama 3, and Gemini 1.5 were 0.4 (SD 0.7), 0.7 (SD 0.76), and 0.33 (SD 0.56) respectively. The combined p-value of GPT 4, Llama 3, and Gemini 1.5 were 1, 1, 0.979 respectively denoting that there was no significant association between the differential diagnosis generated by the LLMs with those generated by the medical experts.

We also compared the LLMs between themselves. The mean Jaccard index of GPT 4 vs Llama 3, GPT 4 vs Gemini 1.5, and Llama 3 vs Gemini were 0.34 (SD 0.18), 0.35 (SD 0.15), and 0.23 (SD 0.12) respectively. The mean Kendall’s Tau of GPT 4 vs Llama 3, GPT 4 vs Gemini 1.5, and Llama 3 vs Gemini were 0.48 (SD 0.66), 0.54 (SD 0.5), and 0.4 (SD 0.66) respectively. The combined p of GPT 4 vs Llama 3, GPT 4 vs Gemini 1.5, and Llama 3 vs Gemini 1.5 were 1, 0.281, and 1 respectively.

We also attempted to find correlation between number of differential diagnoses and Jaccard index to determine whether performance was dependent on the number of differential diagnoses. However no significant correlation was found for GPT 4 (p=0.061), Llama 3 (p=0.091), and Gemini (p=0.355). Similarly, we did not find any significant correlation between number of differential diagnosis and Kendall’s Tau for GPT 4 (p=0.058), Llama3 (p=0.341), and Gemini 1.5 (p=0.404).

The summary of the comparison is shown in table 1.

**Table 1:** Comparison between the medical experts and the large language models

|  | Jaccard Index | Kendall’s Tau | p-value |
| --- | --- | --- | --- |
| Expert vs GPT 4 | 0.30±0.17 | 0.19±0.70 | 1 |
| Expert vs Llama 3 | 0.21±0.13 | 0.40±0.76 | 1 |
| Expert vs Gemini 1.5 | 0.24±0.11 | 0.33±0.56 | 0.979 |
| GPT 4 vs Llama 3 | 0.34±0.18 | 0.48±0.66 | 1 |
| GPT 4 vs Gemini 1.5 | 0.35±0.15 | 0.54±0.50 | 0.281 |
| Llama 3 vs Gemini 1.5 | 0.23±0.12 | 0.40±0.66 | 1 |

## Discussion

The results of this study offer a critical insight into the differential diagnosis capabilities of LLMs in the field of infectious diseases. Notably, the performance of GPT 4, Llama 3, and Gemini 1.5 varied significantly, both in comparison with expert-generated diagnoses and amongst each other, highlighting the difficulties of integrating AI in medical diagnostics.

Firstly, the quantity of differential diagnoses generated by the LLMs differed from that of the medical experts, with Gemini 1.5 producing more diagnoses on average, and Llama 3 the fewest. This suggests that different models may have varying thresholds for diagnostic inclusivity. Gemini 1.5’s higher count could indicate a broader but potentially less precise diagnostic approach, whereas Llama 3’s conservative output might reflect a more focused but potentially under-inclusive diagnostic strategy. GPT 4’s performance sat between these two extremes, suggesting a more balanced approach. These differences underscore the need for tailored calibration of AI tools to align with the desired clinical specificity and sensitivity.

The Jaccard index and Kendall’s Tau scores provide a quantitative measure of the similarity between the AI-generated diagnoses and those of human experts. The scores were modest across all models, indicating that while AI can potentially mimic expert diagnostic patterns to some extent, there remains a gap in achieving medical expert level accuracy and reliability. This gap is further reflected in the non-significant p-values associated with these comparisons, suggesting that the AI models could not significantly match the expert diagnostic decision-making processes.

The lack of significant correlation between the number of differential diagnoses and both the Jaccard index and Kendall’s Tau for each model suggests that increasing number of potential diagnoses does not necessarily affect diagnostic accuracy or alignment with expert opinion. This finding is pivotal for AI development, emphasizing the need for improving the quality of diagnoses rather than the quantity.

Given the rapid evolution and integration of AI in healthcare, these findings suggest several future directions:

1. Model Training and Validation: There is a clear need for further training of these models, using diverse and comprehensive datasets that can better replicate the complex decision-making processes of medical experts.
2. Customization and Configuration: AI models might benefit from being customizable to specific clinical settings or diseases, allowing them to be finetuned according to the specific requirements of the task. Epidemiological considerations should also be taken into account while generating differential diagnosis.
3. Integration with Clinical Workflows: Implementing these AI tools in real clinical workflows will require careful consideration of their limitations and strengths, ensuring they complement rather than complicate the diagnostic process.
4. Ethical and Regulatory Considerations: As AI tools become more prevalent in clinical settings, it will be critical to consider the ethical implications of their use, particularly in terms of accountability, transparency, and the potential for bias.

Our study had several limitations including its reliance on simulated cases, which may not fully capture the complexity and variability encountered in real-world clinical scenarios. While simulated cases allow for a controlled comparison of LLMs, they inherently lack the unpredictability and nuanced details often present in actual patient interactions. Additionally, the study’s focus on a limited number of infectious diseases might not generalize across the broader spectrum of conditions that these LLMs might encounter in practical settings. Furthermore, the study only evaluates the performance of three specific LLMs, which may not represent the capabilities of other emerging models or different configurations of similar technologies.

While the potential of AI in augmenting infectious disease diagnostics is evident, the effectiveness of these tools in real-world settings remains conditional on significant improvements in accuracy, reliability, and integration. The path forward should be guided by continuous collaboration between AI developers, clinicians, and regulatory bodies to ensure that these tools enhance, rather than hinder, medical diagnostics and patient care.

## Conclusion

This study highlights the evolving capabilities and current limitations of LLMs in generating differential diagnoses for infectious diseases. While GPT 4, Llama 3, and Gemini 1.5 demonstrate varying degrees of effectiveness in replicating expert-level diagnostic accuracy, none achieved significant alignment with expert-generated diagnoses, indicating substantial room for improvement. These findings emphasize the necessity for continued development and refinement of AI technologies in healthcare to enhance their practical utility and reliability. Ideally matching should be done with real cases instead of simulated ones with proper ethical considerations. Moving forward, it will be crucial to bridge the gap between AI potential and clinical application through rigorous validation, ethical considerations, and seamless integration into existing medical workflows, ensuring that these tools genuinely enhance patient outcomes and healthcare delivery.

## Data Availability

All data produced in the present study are available upon reasonable request to the authors

## Disclosure on writing assistance

Writing assistance was obtained from GPT 4. However, the authors are solely responsible for the content of the article.

## References

1. Li C, Ye G, Jiang Y, Wang Z, Yu H, and Yang M. Artificial Intelligence in Battling Infectious Diseases: A Transformative Role. Journal of Medical Virology. 2024 Jan; 96:e29355. doi: 10.1002/jmv.29355

2. Tzeng IS, Hsieh PC, Su WL, Hsieh TH, and Chang SC. Artificial Intelligence-Assisted Chest X-ray for the Diagnosis of COVID-19: A Systematic Review and Meta-Analysis. Diagnostics. 2023 Feb 5; 13:584. doi: 10.3390/diagnostics13040584

3. Xin Y, Li H, Zhou Y, Yang Q, Mu W, Xiao H, Zhuo Z, Liu H, Wang H, Qu X, Wang C, Liu H, and Yu K. The Accuracy of Artificial Intelligence in Predicting COVID-19 Patient Mortality: A Systematic Review and Meta-Analysis. BMC Med Inform Decis Mak. 2023 Aug 9; 23:155. doi: 10.1186/s12911-023-02256-7

4. Tran NK, Kretsch C, LaValley C, and Rashidi HH. Machine Learning and Artificial Intelligence for the Diagnosis of Infectious Diseases in Immunocompromised Patients. Current Opinion in Infectious Diseases. 2023 Aug; 36:235– 42. doi: 10.1097/QCO.0000000000000935

5. Giacobbe DR, Zhang Y, and De La Fuente J. Explainable Artificial Intelligence and Machine Learning: Novel Approaches to Face Infectious Diseases Challenges. Annals of Medicine. 2023 Dec 12; 55:2286336. doi: 10.1080/07853890.2023.2286336

6. Smith K and Kirby J. Image Analysis and Artificial Intelligence in Infectious Disease Diagnostics. Clinical Microbiology and Infection. 2020 Oct; 26:1318– 23. doi: 10.1016/j.cmi.2020.03.012

7. Kim JI, Maguire F, Tsang KK, Gouliouris T, Peacock SJ, McAllister TA, McArthur AG, and Beiko RG. Machine Learning for Antimicrobial Resistance Prediction: Current Practice, Limitations, and Clinical Perspective. Clin Microbiol Rev. 2022 Sep 21; 35:e00179–21. doi: 10.1128/cmr.00179-21

